# Validation of the Kuwait Progression Indicator Score for predicting progression of severity in COVID19

**DOI:** 10.1101/2020.05.21.20108639

**Authors:** Sarah Al Youha, Suhail A. Doi, Mohammad H. Jamal, Sulaiman Almazeedi, Mohannad Al Haddad, Mohammad AlSeaidan, Ali Al-Muhaini, Fahad Al-Ghimlas, Salman Al-Sabah

## Abstract

**Introduction:** Identifying patients with COVID-19, at risk of having a severe clinical course during their hospitalization is important for appropriate allocation of clinical resources. We recently described the ‘Kuwait Progression Indicator’ based on laboratory findings, in an initial training cohort derived from the first series of 1096 consecutive patients admitted to Jaber Al-Ahmad Al-Sabah Hospital in Kuwait. The aim of this study was to validate the KPI scoring system in an independent cohort of patients with COVID-19.

**Methodology:** Data was collected prospectively for consecutive patients admitted to Jaber Al-Ahmad Al-Sabah Hospital in Kuwait between 24^th^ February – 28^th^ April 2020. Patients were grouped according to the severity of their clinical course as their main outcome, based on clinical and radiological parameters, with ICU admission and death as secondary outcomes. Model discrimination was assessed through the area under the receiver operating characteristic curve (AUC) while model calibration was assessed through a calibration plot and measures of slope and calibration in the large (CITL).

**Results:** Of 752 patients not used in model development previously, 414 met the criteria for inclusion in this validation study. The baseline characteristics for these 752 patients were similar to the patients that were included in our validation cohort. The area under the curve was equal to 0.904 (95% CI, 0.867-0.942), indicating good model discrimination. The calibration plot and CITL confirmed reasonably good model calibration. Sensitivity and specificity were above 90% for the low and high risk levels respectively.

**Conclusions:** We were able to validate our previously described laboratory based prognostic scoring system for COVID-19 patients, to predict which patients progressed to a severe clinical course.

## INTRODUCTION

As the number of COVID-19 cases continues to rise around the world, lockdown measures are becoming more challenging to maintain [1]. Adapting to life with SARS-CoV-2 in our midst is rapidly becoming an option that countries are starting to exercise [2]. Appropriate triage and rationing of medical supplies are becoming even more important as a result. Moving forwards, prognostic clinical scoring systems for COVID-19 will become valuable tools, as they can potentially allow early identification of individuals who are at high risk of poor outcomes and require intensive clinical resources. These high-risk individuals may also be candidates for earlier, more aggressive interventions [3,4].

A recent systematic analysis by Wynants et al.[5] evaluated several COVID-19 prediction models and deemed most of them to be poorly reported and at high risk of bias. A large majority of the reviewed models were not performed in consecutive cohorts of patients and were not validated. In addition, most of them were based on radiological findings, such as CT scans, which are not practical to perform for everyone on initial presentation.

We recently described a prognostic predictive scoring system, the ‘Kuwait Prognostic Indicator’ (KPI) [6]. This scoring system was based on laboratory findings in consecutive patients admitted to Jaber Al-Ahmad Al-Sabah Hospital in Kuwait. This cohort was composed of patients with a wide spectrum of clinical presentations, who received identical investigations and treatment protocols, as they were all admitted to a single site. This is due to the fact that prior to the first COVID-19 case even being diagnosed in Kuwait, the Kuwaiti government initiated COVID-19 screening for all travelers from problem areas abroad and placed them in institutional quarantine in Jaber Al-Ahmad Al-Sabah Hospital, irrespective of symptomology[7].

The aim of this study was to validate our clinical scoring system, the KPI, in an independent group of patients. Our hypothesis was that this scoring system would be valid when applied to a different patient cohort.

## METHODS

### Study Design

We obtained ethical approval from the Kuwait Ministry of Health ethical review committee. This study is a validation study that continued data collection of consecutive patients in a previous prospective cohort study that defined the key predictors of COVID19 severity and created a laboratory test based score at diagnosis for predicting progression of COVID19 to more severe illness. The latter was called the Kuwait Progression Indicator (KPI) score and was developed amongst patients admitted to Jaber Al-Ahmad Al-Sabah Hospital in Kuwait between 24 February 2020 and 20 April 2020 [6].

The definition of cases and inclusion criteria were the same as previously reported [6]. The training cohort in the original study comprised of 700/1096 patients of whom 406 had reached an outcome by the end of the study and formed the cohort for development of the KPI score. We now exclude these 406 subjects and report the external validation of the KPI score on an expanded cohort of 752 patients that form this independent validation cohort. As with the development cohort, the same algorithmic severity grouping [6] was the main outcome with ICU admission and death being secondary outcomes.

### Statistical Analysis

The scores were generated for the patients as per the KPI scoring system depicted in Table 1. We present categorical variables summarized using percentages and continuous variables summarized using medians with interquartile ranges. Association of scores with documented hard endpoints (ICU admission or death) were assessed using binary logistic regression.

**Table 1.** The KPI score

| **Kuwait progression Indicator score (KPI score) for COVID19**<br>**Please give your patient zero points if criterion not met** |  |  |
| --- | --- | --- |
| Criterion | Points | Your patient |
| Age ≥ 41 years | 4 | |
| CRP ≥ 7 mg/L | 2 | |
| Procalcitonin ≥ 0.05 ng/ml | 16 | |
| Lymphocyte percent ≥ 31.5% | -9 | |
| Monocyte percent ≥ 9.2% | -8 | |
| Albumin ≥ 39.5 g/L | -15 | |
| **TOTAL** |  |  |
| **LOW PROGRESSION RISK TOTAL ≤ -7** |  |  |
| **HIGH PROGRESSION RISK TOTAL ≥ 16** |  |  |
| **UNCERTAIN PROGRESSION RISK TOTAL 8 TO 15** |  |  |

The discrimination of the model for those who had reached an outcome was evaluated through assessment of the area under the receiver operating characteristic curve (C statistic). Operating characteristics of the score were assessed using interval likelihood ration given that the KPI has three levels of risk. Calibration of the model was assessed using pmcalplot in Stata [8] reporting calibration slope and calibration in the large (CITL). Calibration-in-the-large (CITL) indicates the difference between the observed prevalence and the mean predicted probability, which should be zero. This population-based level does not guarantee calibrated risks at the individual patient level [9]. The calibration slope indicates whether the model coefficients are underfitted (slope > 1) or overfitted (slope < 1). Thus the calibration slope expresses predictions that are too extreme or not extreme enough. All analyses were performed using Stata MP version 15 (College Station, TX, USA) and the confidence level was set at 95%.

## RESULTS

### Baseline Characteristics

Of these 752 patients, 690 were updated from the original cohort and 62 new patients added till 28 April 2020. Males were represented similarly to the development study (649/752; 86%) and comorbidities were more represented in this cohort (225/752; 30%). Their mean age was the same as the development cohort (41.7 years; range, 1 – 87 years).

Of the 752 patients, clinical course could be defined based on the algorithm [6] in 414 patients (the rest had not yet reached the outcome) and these formed the cohort for the model validation. Details of the basic characteristics of these patients are given in Table 2. Of note, the final KPI score could be computed (missing data) for 691/752 patients (92%). The range of KPI scores was from −32 to 22. Of the 414 patients with outcome information, the clinical course was mild in 324 (78%) and mod-severe in 90 (22%). Of these 90 severe cases, 77 had been admitted to the intensive care unit because of a worsening respiratory status and 30 had died.

**Table 2:** Baseline Characteristics of patients with COVID-19

| Characteristic | Median (IQR) or<br>N (%)<br>With clinical<br>course data<br>(N=414) | Median<br>(IQR) or<br>N (%)<br>All data<br>(N=752) |
| --- | --- | --- |
| Age | 44 (33 - 55) | 40 (32 – 50) |
| Males | 466 (72.5%) | 888 (81.0%) |
| Nationality |  |  |
| Asian* | 209 (50.5%) | 494 (65.7%) |
| Kuwaiti | 128 (30.9%) | 141 (18.7%) |
| Others | 77 (18.6%) | 117 (15.6%) |
| Duration of stay<br>(days)** | 13 (8 - 18) | 2 (6 – 13) |
| Diabetes Mellitus | 79 (19.1%) | 106 (14.1%) |
| Hypertension | 89 (21.5%) | 113 (15.0%) |
| Asthma | 25 (6.0%) | 29 (3.9%) |
| CAD/IHD*** | 23 (5.6%) | 25 (3.3%) |
| ICU admission | 77 (18.6%) | 77 (10.2%) |
| Death | 30 (7.2%) | 30 (4.0%) |

\*India/Bangladesh/Philippines
\*\* Shorter stay for the whole cohort as included those more recently admitted whose course was yet to be determined
\*\*\*Coronary/ischaemic heart disease

### Validation of the KPI score

The area under the curve (AUC) was equal to 0.904 (95% CI, 0.867–0.942), which indicates good model discrimination (figure 1). A calibration plot of observed against expected probabilities for assessment of prediction model performance demonstrated reasonably good model calibration (Figure 2).

**Figure 1.**
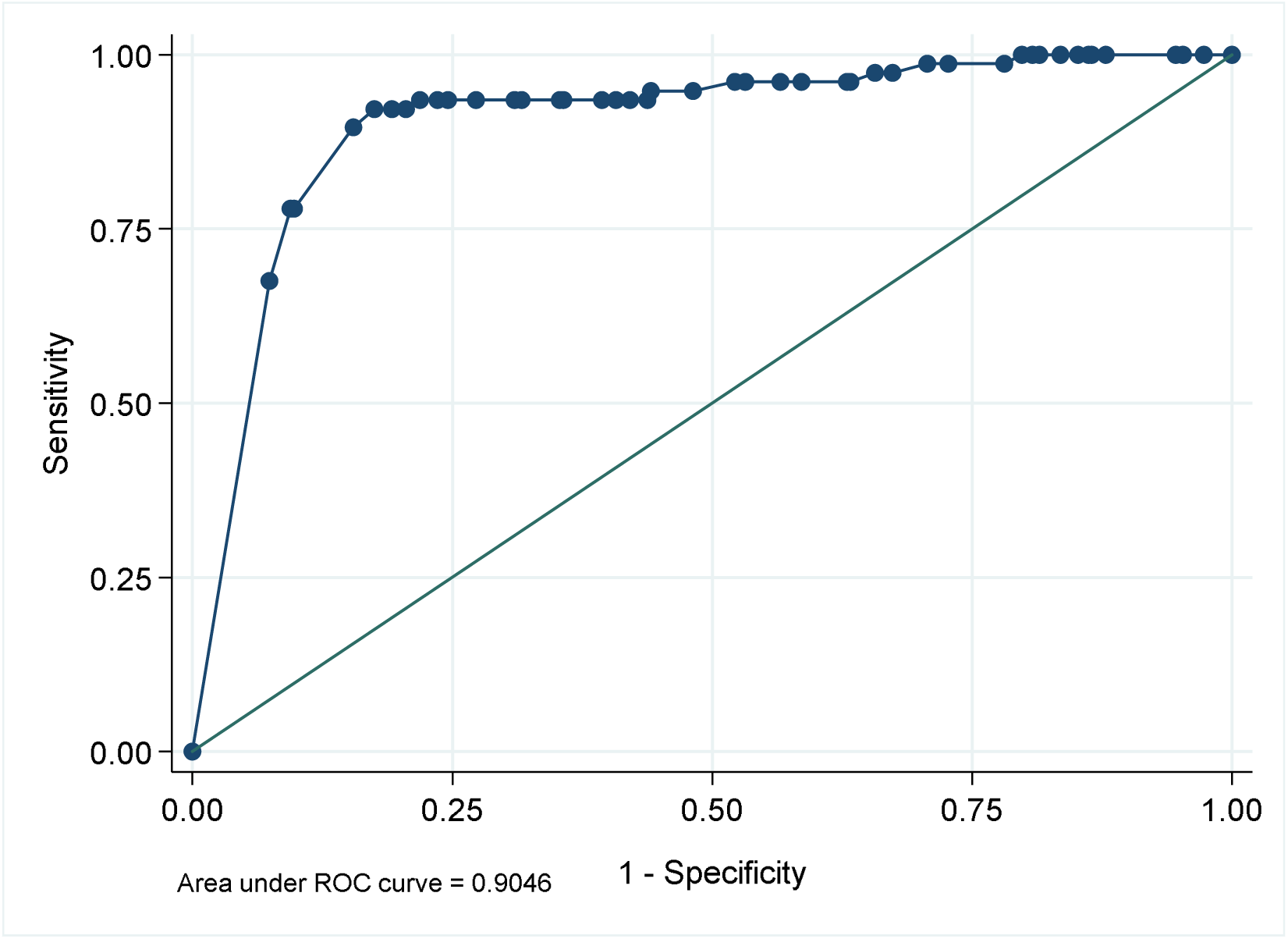
Discrimination of the KPI score depicted via a ROC curve. The area under the curve (AUC) is 0.90 suggesting good discrimination.

**Figure 2.**
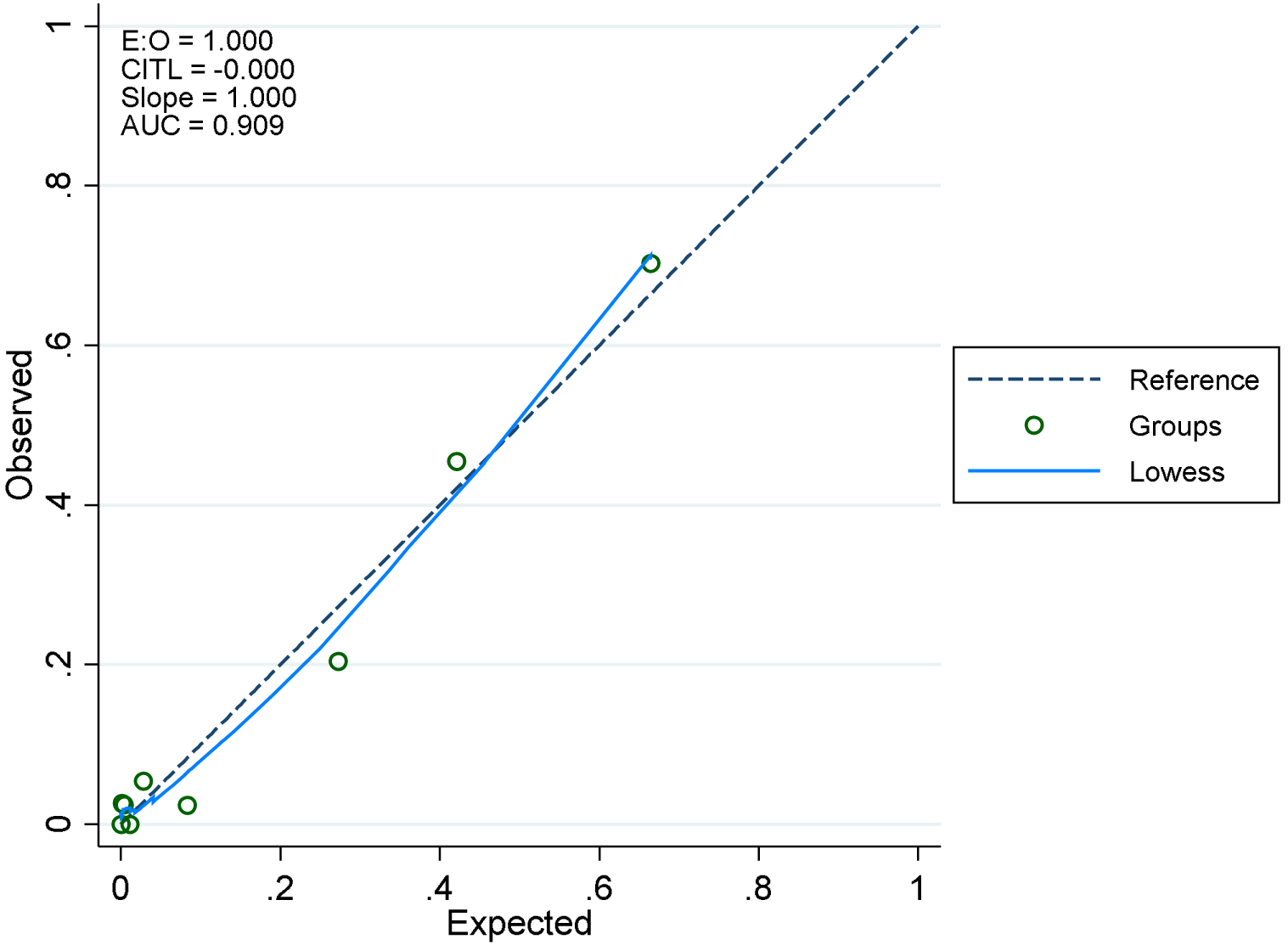
Calibration plot depicting the calibration of the KPI score. CITL stands for calibration-in-the-large.

The risk groups also demonstrated good discrimination of the various outcomes (Table 3). At a cut-off at the high-risk level, the sensitivity was 77.9% (95% CI 67.0 – 86.6%) and specificity was 90.2% (95% CI 86.3 – 93.4%) for a severe status. The likelihood ratio for a high-risk level was 7.98 (95% CI 5.54 – 11.50). Similarly, at a cut-off at the low risk level, the sensitivity was 96.1% (95% CI 89.0 – 99.2%) and the specificity was 46.8% (95% CI 41.0 – 52.7%) for a severe status. The likelihood ratio for a low risk level was 0.08 (95% CI 0.03 – 0.25). Thus, a patient at the low risk level practically rules out while a patient at the high-risk level practically confirms likely progression of COVID19 in terms of severity that requires hospital treatment.

**Table 3:**
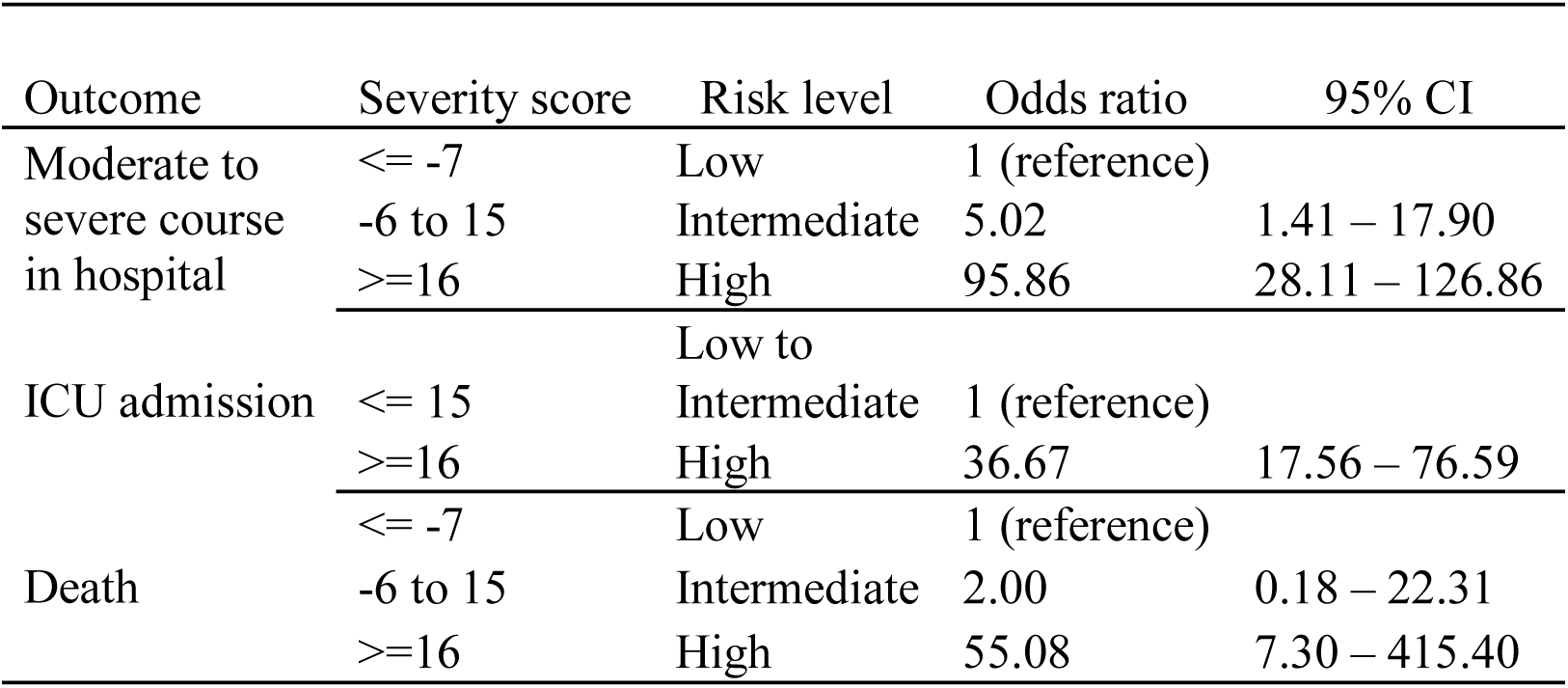
Odds ratio of outcomes by category of severity score

## DISCUSSION

We were able to validate our laboratory based prognostic scoring system for COVID-19 patients, the KPI, in an independent patient cohort, to predict which patients progressed to a severe status. Based on their KPI score, we managed to stratify patients to being ‘low’, ‘uncertain’, or ‘high’ progression risk. Namely, a KPI score greater or equal to 16 was predictive of high risk of progression to a moderate to severe course in the hospital, sometimes requiring ICU admission and/or leading on to death. We showed that our model had good discrimination (AUC = 0.904, compared to AUC = 0.83 in the cohort used to develop our model [6]) and we were able to calibrate the prediction model performance, demonstrating that it was reasonably good.

This scoring system can be used clinically to identify patients on admission, who are likely to progress to a moderate to severe clinical course during their hospitalization period. By early identification of these patients, resources can be mobilized earlier, and physicians may consider instigating more aggressive therapeutic interventions prior to severe clinical deterioration. This predictive tool may also be a useful adjunct for clinicians, equipping them with means of providing patients and their families with more information, when counselling them regarding their expected clinical course in hospital [5].

The laboratory parameters utilized in our cohort are simple and can easily be performed for patients on admission to the hospital. They are based on non-specific biochemical inflammatory and hematological measures, that have been shown to be independently associated with poor clinical outcomes for COVID-19 patients in other studies [10–15]. The KPI scoring system is essentially, an aggregate measure of these serological changes, categorized via a machine learning algorithm to enable them to be utilized to make clinical prognostic predictions for patients with COVID-19. Age was also another independent predictor incorporated in our model that was shown by other authors to be an important risk factor for unfavorable disease progression [16,17].

Our model had several advantages, compared to other prognostic models for COVID-19 severity [5]. It was based on laboratory investigations, which are objective quantifiable measures, compared to scoring systems that rely on self-reported symptoms and other subjective parameters. Our cohort was based on consecutive patients, mitigating issues with selection bias. A significant proportion of our study subjects were also asymptomatic, making our hospitalized patient sample more heterogenous in clinical severity on initial presentation compared to other cohorts used to develop prognostic scoring systems described in the literature [4,5].

Our limitations include that the number of patients in our validation cohort was relatively modest. Another consideration is that our validation was performed in the same country, in Kuwait. Future work will aim to validate our model in other centers, in different countries. We also plan to make our scoring system available online for practicing clinicians to make it more accessible and easier to use.

## Data Availability

All relevant data included in the manuscript is available on request

